# Molecular Identification of *Staphylococcus aureus* Complex Strains and Evaluation of Methicillin Resistance Among Human Community Isolates from Biological Samples at Angré Hospital University in Abidjan, Côte d’Ivoire

**DOI:** 10.1101/2025.06.02.25328779

**Authors:** DNC Obouayeba, S Niemann, GA Bahan, GE Coulibaly, KAA Kofi, YM Coulibaly-Diallo, AJ Djaman, A Kacou-Ndouba

## Abstract

The *Staphylococcus aureus* complex is a group of bacterial species that share several biological and pathogenic characteristics. It mainly includes *Staphylococcus aureus*, as well as closely related species such as *Staphylococcus argenteus* and *Staphylococcus schweitzeri. S. schweitzeri* is the only species in this complex that predominantly occurs in African fauna. However, data on a possible transmission of *S. schweitzeri* to humans are still limited. From a phenotypic perspective, differentiating the species within the *S. aureus* complex remains challenging, which may obscure the true diversity of the complex. As a result, strains identified as *S. aureus* might belong to other species. This study aimed to determine the distribution of *S. aureus* complex species and the methicillin resistance rate among clinical isolates collected in Abidjan. **Methods:** We analyzed 330 clinical strains previously identified as *S. aureus* using conventional phenotypic methods. This was supplemented by conventional PCR assays targeting the *nuc* gene and the NRPS gene, which allows discrimination among members of the complex. Methicillin-resistant *S. aureus* (MRSA) was detected using the cefoxitin disk diffusion test, and the presence of the *mecA* gene was assessed by PCR in MRSA-positive isolates. **Results:** All isolates tested positive for the *nuc* gene (270 bp), and 96.97% were positive for the NRPS gene (160 bp). *S. aureus* was confirmed in 317 isolates (>96%). MRSA accounted for 50.30% of all isolates, with a statistically significant increase in MRSA prevalence with age and department hospital (p = 0.00049, P=0.0001857). **Conclusion:** PCR detection of the *nuc* and NRPS genes confirmed that nearly all isolates belonged to the *S. aureus* species. Therefore, our data suggest that *S. schweitzeri* has not yet been able to cross the species barrier to humans in Côte d’Ivoire. Over half of the biological samples from the university hospital contained methicillin-resistant *S. aureus* strains, highlighting a significant public health concern.

**Author summary:** *S. aureus* belongs to the *S. aureus* complex, along with, for example, *S. schweitzeri* and *S. argenteus*. While much is already known about *S. aureus*, knowledge about the pathogenicity and spread of *S. schweitzeri*, which has so far mainly been found in monkeys and fruit bats in sub-Saharan Africa, is still very limited. It is not yet known whether it is a zoonotic pathogen. This is the first study in Côte d’Ivoire in which molecular tools were used to differentiate *S. aureus* from other closely related species in clinical samples collected from a hospital in Abidjan, the capital city. Two genes, *nuc* and NRPS, were targeted to determine the species present. We confirmed that almost all of our samples were positive for *S. aureus* and negative for *S. schweitzeri*. This suggests that *S. schweitzeri* has not yet evolved the ability to transmit from animals to humans and cause infection. However, more than half of the bacteria were resistant to the important antibiotic methicillin, posing a real public health problem. These results highlight the need to improve local surveillance of bacterial infections to enable them to be better understood and treated.

## Introduction

*Staphylococcus aureus* is a ubiquitous bacterium found in the normal flora of humans and animals, as well as in environment, but it can also cause a wide range of infections, from superficial conditions to severe invasive diseases [1]. In 2015, an epidemiological study conducted in Bouake (Côte d’Ivoire) identified *S. aureus* as the main Gram-positive germ isolated from community bloodstream infections [2].

Yet, despite this constant presence in clinical practice, very few studies have been published on this pathogen since then. This lack of local data contrasts sharply with the situation in high-income countries, while in sub-Saharan Africa, the burden of community-based invasive *S. aureus* infections remains high [3].

At the medical biology laboratory of the University Hospital Center (CHU) Angré (Abidjan, Côte d’Ivoire), *S. aureus* remains one of the most frequently isolated pathogens in community infections. Internal records show that it is the most isolated Gram-positive germ and the second most common bacterium after *Escherichia coli* in community bloodstream infections. About a hundred cases from blood cultures are recorded on average each year.

In sub-Saharan Africa, infectious disease management still faces a deficit in diagnostic infrastructure and a lack of local data. Apart from a few well-monitored pathogens such as *Plasmodium* spp., or *HIV*, the majority of bacterial infections remain outside the specific monitoring devices and are generally treated empirically, without real diagnostic confirmation [4]. This approach contributes to an underestimation of the real burden of these infections in these countries.

Several authors had drawn attention to this lack of recognition. Although not included on the official list of neglected tropical diseases, serious community-acquired bacterial infections should be considered as such, due to their real impact on public health and their low visibility in research priorities [5]. This lack of interest often results in a direct and sometimes inadequate transfer of data produced in industrialized countries to tropical contexts, without really taking into account the realities on the ground.

In this context, *S. aureus* infections are a good example of this discreet but persistent form of neglect. It is a common pathogen, well known, but paradoxically still poorly documented in countries with limited resources. It is a good example of these infections that could be described as “neglected diseases of a new kind,” not because they are completely ignored, but because they do not receive the attention they deserve in research programs, especially in Africa.

This lack of interest is all the more worrying as our understanding of *S. aureus* has evolved a lot in recent years. What was long thought to be a single species is actually part of a larger genetically related group: the *Staphylococcus aureus* complex, which also includes *Staphylococcus argenteus* and *Staphylococcus schweitzeri, Staphylococcus roterodami,* and *Staphylococcus singaporensis* [6]. This genetic diversity calls into question certain certainties, particularly in terms of diagnosis in areas where these species are still little studied.

Among these, *S. schweitzeri* is considered an emerging pathogen and is the only species in the complex primarily found in African fauna [7], particularly in bats and primates. Although *in vitro* data indicate that *S. schweitzeri* might be as virulent as *S. aureus* [8], the bacterium has rarely been detected in humans to date. The few documented cases in humans usually involve colonisation of the nasopharyngeal cavity, which may be related to contact with wild animals [9]. However, the identification of *S. aureus* is typically based on phenotypic criteria, which can lead to misclassification due to the strong biochemical and microbiological similarities between species. As a result, some strains suspected to be *S. aureus* may actually belong to other species within the *S. aureus* complex. In this context, specific molecular tools are required for the accurate identification of species within the complex. PCR targeting the *S. aureus*-specific thermostable nuclease gene (*nuc*) and the non-ribosomal peptide synthetase (NRPS) gene are particularly useful for this purpose [10, 11]. The *nuc* gene serves as a genetic marker for *S. aureus*, while *S. schweitzeri* carries a divergent nuclease gene (*nucM*), whose sequence shows variations that prevent amplification by the primers described by Brakstad et al. [12]. Consequently, in this assay, *S. schweitzeri* appears *nuc-*negative, whereas *S. aureus* and *S. argenteus* are *nuc*-positive [13].

Additionally, analysis of the NRPS gene allows differentiation of species within the complex based on the size of the PCR amplicon. *S. aureus* produces a 160 bp fragment, while *S. schweitzeri* and *S. argenteus* generate a 340 bp fragment [11]. This approach helps to avoid confusion with *S. aureus* in clinical settings.

Our understanding of *S. schweitzeri* as well as *S. argenteus* as emerging pathogens is still poor. Due to its close relationship to *S. aureus*, it is possible that both *S. schweitzeri* and *S. argenteus* have been misidentified as *S. aureus* in humans. If *S. schweitzeri* could be transmitted from animals to humans, contact with animals colonised with *S. schweitzeri* or their feces (e.g. from fruit bats) could pose a risk for zoonotic infections.

In this study, the first of its kind in Côte d’Ivoire, we employed PCR targeting the *nuc* and NRPS genes to characterize strains isolated from biological samples of human infections collected at Angré University Hospital, aiming to evaluate the diversity of *S. aureus* complex species in this population. Furthermore, this study provides novel data on the prevalence of MRSA in clinical isolates in Côte d’Ivoire.

## Material and methods

### Study site and sample collection

This study was conducted at the medical biology laboratory (bacteriology unit) of the University Hospital Center (CHU) of Angré in Abidjan, Côte d’Ivoire. Bacterial isolates were obtained from various biological samples collected from both inpatients and outpatients between December 2021 and February 2024.

### Bacterial Isolation and Culture

The isolates used in this study were recovered from strains preserved either in deep nutrient agar at room temperature or frozen at –80 °C in brain heart infusion broth supplemented with 10% glycerol. These strains, stored between December 2021 and February 2024, were reactivated prior to analysis and cultured on nutrient agar and Chapman agar for further testing. The reference strain *S. schweitzeri* (FSA084^T^), isolated from a red-tailed monkey in Gabon [ 7] and described by Tong et al [14], was provided by the Institute of Medical Microbiology, University Hospital of Münster, Germany. *S. aureus* strains ATCC 43300 and ATCC 25923, from the medical biology laboratory of the Angré University Hospital, were used for comparison purposes

## Identification of isolates

### Phenotypic Identification

Isolates of the *S. aureus* complex were first identified by classical phenotypic microbiological methods, including Gram staining, catalase, DNase and free coagulase assays. Identification was also done by automated methods using the compact Vitek 2 system and the Gram-Positive (GP) color card (BioMérieux, Marcy-l’Étoile, France).

## Genotypic Identification

### Genomic DNA extraction

Genomic DNA was extracted using the Biofact kit (DNA/RNA Amplification, Korea), following the manufacturer’s instructions. Bacterial colonies grown on nutrient agar for 18 to 24 hours were collected, washed, and concentrated in 500 μL of PBS by centrifugation at 8000 g for 10 minutes. The resulting pellet was resuspended in 200 μL of DNA/RNA-free solution. The cell suspension was incubated at 37 °C for 15 minutes, followed by homogenization using a high-speed vortex.

After homogenization, the sample was incubated at 56 °C for 20 minutes after the addition of 250 μL of lysis buffer and 20 μL of proteinase K. After incubation, DNA was precipitated and subsequently washed with Wash Buffers 1 and 2 provided in the Biofact kit. Finally, DNA was eluted using the Biofact elution buffer and stored at -20 °C until further use.

### PCR Amplification of nuc and NRPS Genes

PCR amplification was performed in a CFX96 thermal cycler (Bio-Rad) using the conditions outlined below. Reactions were carried out with Solis Master Mix (Solis Biodyne) in a final volume of 22 μL, consisting of 4 μl of Solis 5x Master Mix, 0.75 μL of each primer (10 μM), 3 μL of template DNA, and 13.5 μL of molecular-grade water.

The specific primers used were *nuc*-F and *nuc*-R for the *nuc* gene, generating a 270 bp amplicon [10], and NRPS-F and NRPS-R for the NRPS gene, producing a 160 bp fragment for *S. aureus* and a 340 bp fragment for *S. schweitzeri* and *S. argenteus* [11]. The primers used for the different PCRs and the amplification conditions are listed in tables 1A and 1B.

**Table 1A.**
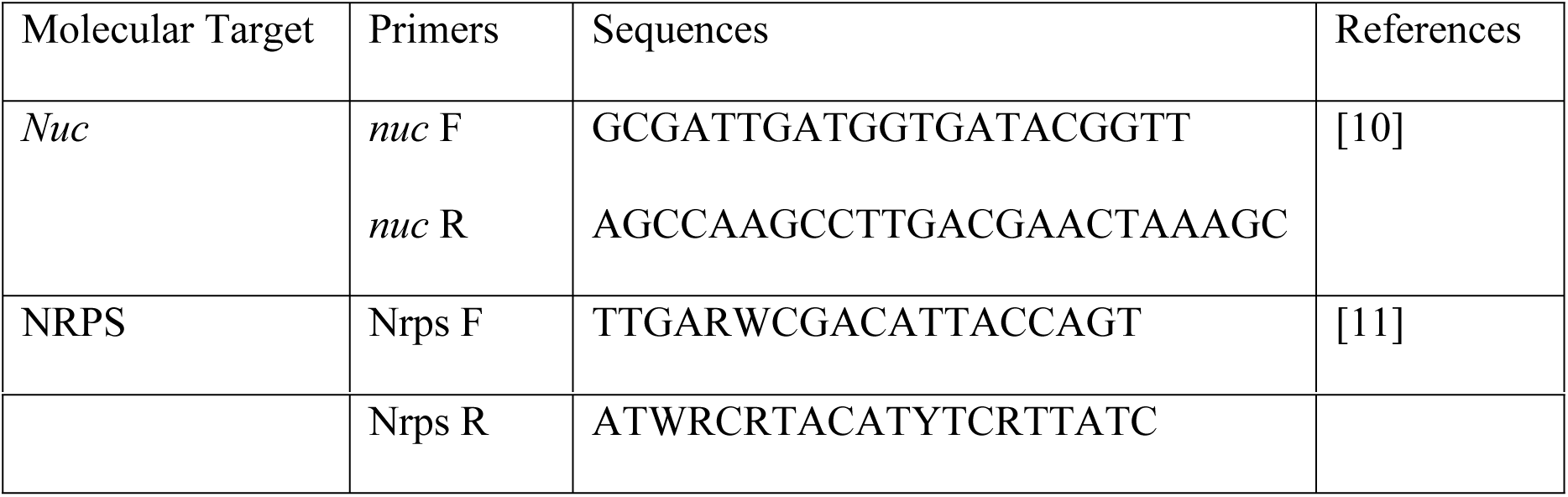
Primer sequences of *nuc* and NRPS genes.

**Table 1B.** *nuc* and NRPS amplification program.

|  | *nuc* gene | NRPS gene |
| --- | --- | --- |
| Initial denaturation | 95°C, 3 min | 94°C, 4 min |
| Denaturation | 95°C, 30 sec | 94°C, 30 sec |
| Hybridization | 59°C, 30 sec | 46°C, 30 sec |
| Elongation | 72°C, 30 s | 72°C, 40 sec |
| Final elongation | 72°C, 10 min | 72°C, 10 min |
| Number of cycles | 30 | 35 |

Amplification conditions included an initial denaturation step, followed by cycles of denaturation, annealing, and elongation. A final elongation step was performed before stopping the PCR for each gene.

Amplification products were visualized by electrophoresis on a 1.5% agarose gel containing SybrSafe® (Invitrogen®)

### Determination of methicillin resistance

The study of methicillin sensitivity was carried out on all isolates. The Mueller-Hinton agar disc scattering method was used with a cefoxitin disc (30 μg). The interpretation of the results followed the recommendations of the Antibiogram Committee of the French Society of Microbiology [15]. An automated method using the Vitek 2 system (BioMérieux) was also implemented to determine antibiotic susceptibility.

Detection of the mecA gene was performed on MRSA strains with primers: *mecA* Foward 5’-TGAGTTGAACCTGGTGAAGTT-3’ and *mecA* Reverse 5’-TGGTATGTGGAAGTTAGATTGG-3’ generating an amplicon of about 855 bp [16], including initial denaturation: 95°C for 5 minutes followed by 35 cycles of Denaturation at 95°C for 30 seconds, hybridization at 58°C for 30 seconds and extension at 72°C for 1 minute, with a final extension at 72°C for 5 minutes. *S. aureus* strains ATCC 43300 (methicillin-resistant *S. aureus* (MRSA)) and ATCC 25923 (methicillin-susceptible *S. aureus* (MSSA)) were used as controls.

### Statistical analysis methods

Data were entered and analyzed using R Software (version 4.3.2) and Microsoft Excel. The Chi-square test and exact Fisher test were used to assess associations and compare proportion as appropriate. A *p* value < 0.05 was considered statistically significant.

### Ethical Considerations

This study was conducted using bacterial strains preserved from clinical biological samples obtained through routine analyses at Angré Hospital university. No patient data, including names, were used in this study, and all information was processed anonymously. This research did not require specific ethical approval.

## Results

### General Specimen and Patient Characteristics

Between December 2021 and March 2024, a total of 330 biological samples were collected in various departments of the Angré University Hospital. All patients resided in urban areas, and no medical history was reported. The mean age of patients was 16.26 ± 23.095 years, ranging from 0 to 78 years.

The distribution of isolates by hospital department was as follows:

- 166 (50.3%) from pediatrics and neonatology,
- 73 (22.1%) of the surgical and traumatology department,
- 49 (15.2%) of the medical department,
- 28 (8.5%) of the intensive respiratory care unit,
- 13 (3.9%) from another service

The majority of samples came from blood samples, which accounted for 74.78% of the samples (247 samples), purulent samples (abscesses, wounds, boils) made up 19.32% of the samples (64), followed by puncture fluids with 4.79% of the samples (16), and finally urine representing 0.78% of the samples (3).

### Phenotypic identification of *S. aureus*

Phenotypic identification of isolates was performed using biochemical assays:

- Catalase: 100% of isolates tested positive
- DNase: 71% of isolates tested positive
- Tube coagulase: 69% of isolates tested positive
- Mannitol fermentation: 67.31% of isolates fermented mannitol

### Genotypic Identification of *S. aureus*

PCR analysis assessed the distribution of *the nuc* and NRPS genes in human infection isolates. All 330 suspected *S. aureus* isolates tested positive for *the nuc* gene (270 bp). The NRPS gene (160 bp) was detected in 96.97% of isolates, while 3.03% of isolates remained negative after several rounds of PCR testing (Figure 1). Thus, 96.97% of isolates were confirmed as belonging to the *S. aureus* species (positive *nuc-PCR* (Figure 2A) and positive NRPS-PCR at 160 bp (Figure 2B).

**Figure 1.**
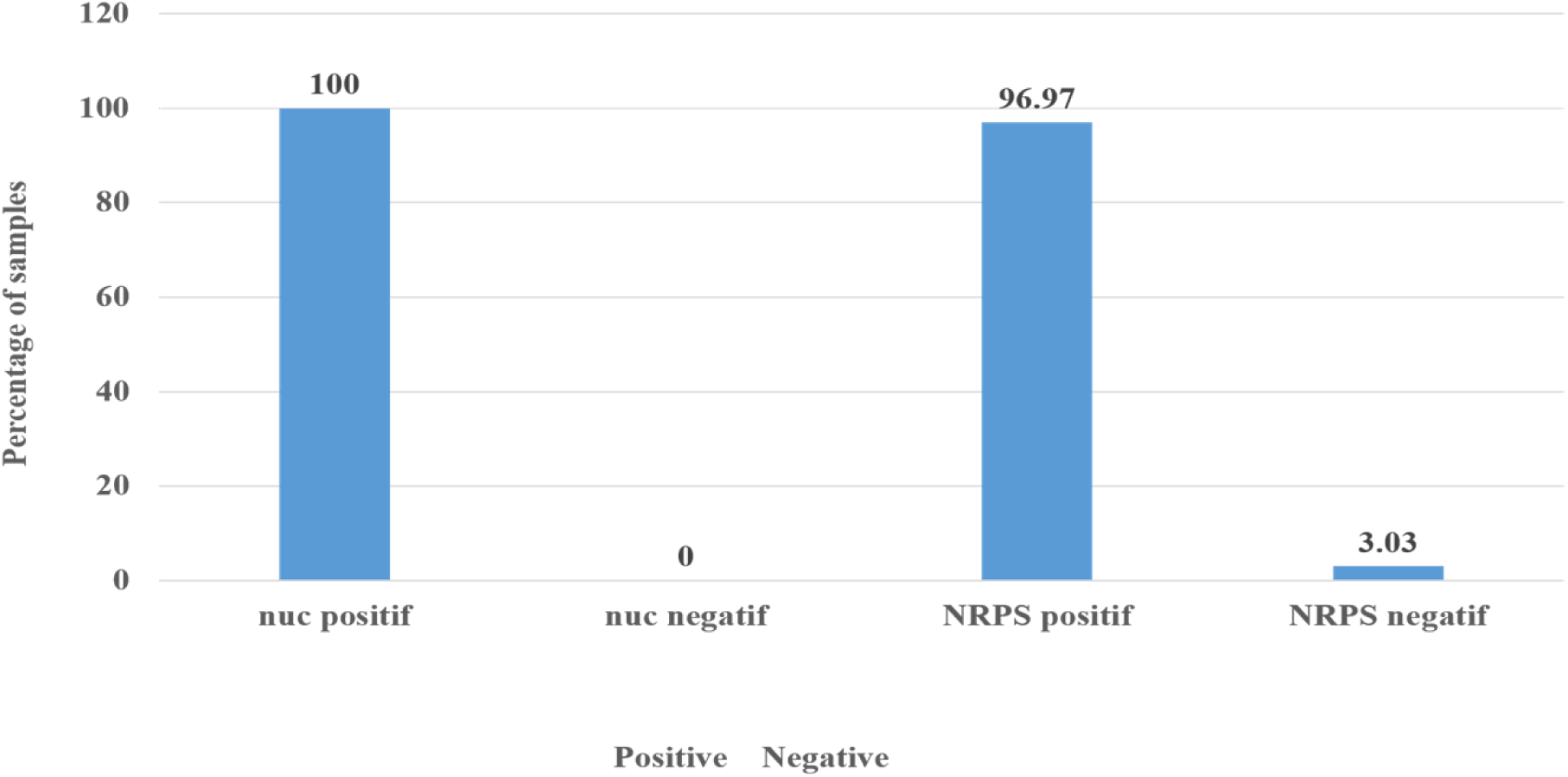
Distribution of the *nuc* and NRPS genes in human infection isolates. Distribution of PCR results for the molecular target *nuc* and NRPS in the 330 clinical isolates. The chart illustrates the frequency of positive and negative detection for each molecular target expressed in percentage.

**Figure 2.**
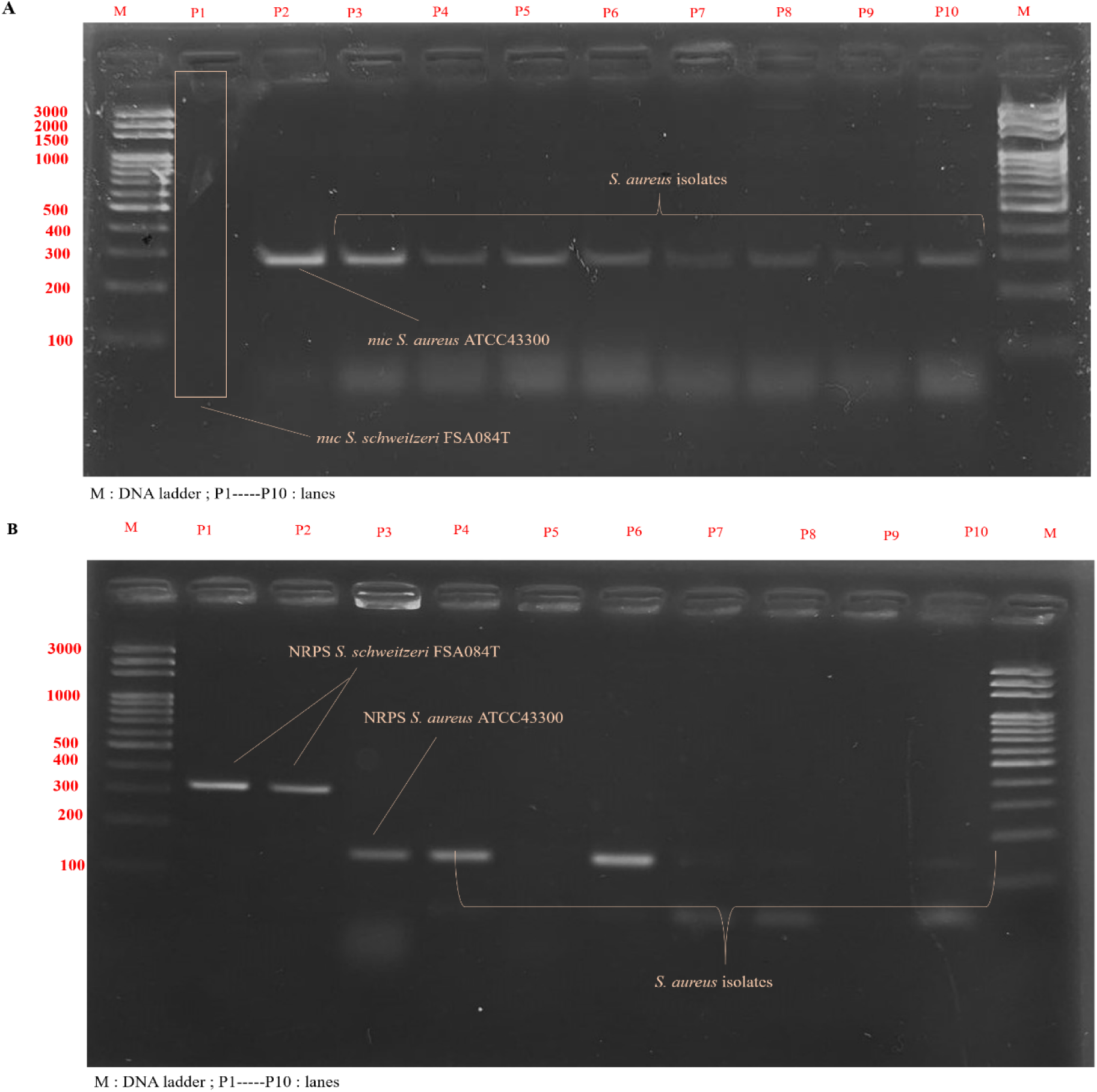
PCR amplification of the *nuc* and NRPS gene in clinical isolates of *S. aureus*. **(A)** Visualization on 1.5% agarose gel under UV light of the *nuc* gene in 8 clinical *S. aureus* isolates. Lane M: molecular weight marker (100 bp DNA ladder); lane 1:*S. schweitzeri* reference strain reference strain FSA084T (negative *nuc*). Lane 2: *S. aureus* reference strain ATCC 43300 (positive control); lanes 3–11: clinical isolates. The presence of a ∼270 bp band indicates a positive result for the *nuc* gene. **(B)** Visualization on 1.5% agarose gel under UV light of the NRPS gene in 7 clinical isolates of *S. aureus*. Lane M: molecular weight marker (100 bp DNA ladder); lanes 1 and 2: *S. schweitzeri* reference strain with a band at approximately 340 bp, indicating a positive result for the *S. schweitzeri* NRPS gene. Lane 3 corresponds to the *S. aureus* reference strain, which shows a band at approximately 160 bp (length of NRPS amplificate from *S. aureus* NRPS). All NRPS-positive isolates produced a band of 160 bp. Lanes 5 and 9 correspond to NRPS-negative isolates. Lanes 7, 8, and 10 show faint or smeared bands, likely due to reduced DNA concentration in these samples

### Methicillin resistance, *mecA* gene distribution and associated factor

Evaluation of methicillin resistance was performed for all isolates among them, 50.3% were resistant to cefoxitin and diagnosed as MRSA, 80.12% were carriers of the *mecA* gene.

The frequency of MRSA was determined and its association with sex, the original department, age and sample type was investigated.

The distribution of *S. aureus* strains varied significantly across hospital departments (Figure 3A, *p* = 0.0001857) as well as across patient age groups (Figure 3B, *p* = 0.00049). The pediatrics and neonatology department had the highest number of isolates, mostly methicillin-susceptible (MSSA), while the surgery and general medicine, geriatrics (adult medicine) departments had a higher proportion of resistant strains (MRSA). Among the 330 samples included, patient age was available for 236 cases; the remaining 94 cases were excluded from the statistical test for association with age but included in the descriptive representations. The peak incidence of MRSA was observed in the age groups and [46–78] years. The distribution of resistant and sensitive strains according to sex and the nature of the biological sample showed no significant variation (Table 2).

**Figure 3.**
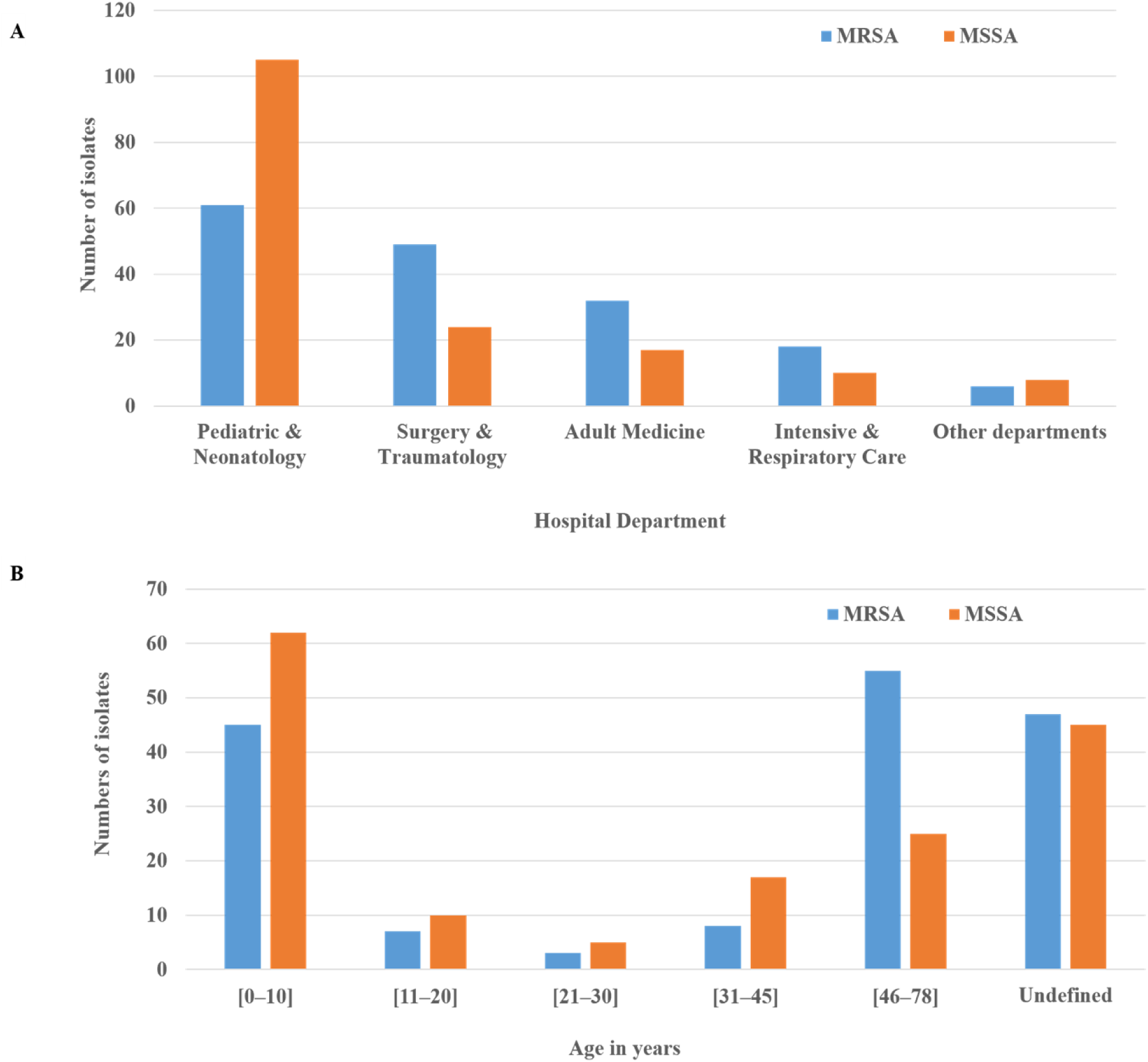
Distribution of *S. aureus* strains according to methicillin-resistance. **(A)** Distribution across hospital departments. MRSA strains predominated in surgery, adult medicine and intensive respiratory care, while MSSA strains were more common in pediatrics and neonatology. **(B)** Distribution of MRSA by age group of patients. Ages are expressed in years. The group marked “undefined” corresponds to patients for whom the age was not reported. This category was not included in the comparative analysis due to missing clinical data.

**Table 2.** Distribution of MRSA and MSSA strains according to biological sample and patient sex.

| Variables studied | Category | MRSA | MSSA | *p* value |
| --- | --- | --- | --- | --- |
| Biological sample | Blood | 117 | 130 | 0.171 |
|  | Pus | 40 | 24 |  |
|  | Puncture Fluids | 8 | 8 |  |
|  | Urines | 1 | 2 |  |
| Sex | Male | 75 | 70 | 0.745 |
|  | Female | 74 | 73 |  |
|  | Undefined | 17 | 21 |  |

Distribution of MRSA and MSSA strains according to biological sample and patient sex. Data are expressed in absolute numbers. *p* values indicate the results of statistical: The Chi-square test was used for large numbers and the Fisher exact test when the numbers were less than 5. These tests evaluating associations between the variables studied. A *p* value less than 0.05 is considered significant.

## DISCUSSION

### Identification of *S. aureus* and absence of *S. schweitzeri*

Identification of *S. aureus* complex strains has traditionally relied on phenotypic criteria, such as colony morphology, coagulase production, and mannitol fermentation. However, these approaches have certain limitations, mainly due to the phenotypic similarity observed among species within the *S. aureus* complex. To overcome these limitations, genotypic identification based on PCR targeting specific genes offers a more reliable and discriminating method.

In this study, amplification of the *nuc* gene, a specific marker of *S. aureus*, yielded positive results in 100% of the strains previously identified phenotypically as *S. aureus*. These findings are consistent with numerous other studies reporting 100% specificity of the *nuc* gene among human *S. aureus* isolates ([17], [18], [19]).

In parallel, analysis of the NRPS gene, another marker associated with, showed positivity in over 96% of the strains, with a 160 bp amplicon characteristic of this species. These results further support the idea that combining multiple molecular markers, such as *nuc* and NRPS, enables more accurate identification of *S. aureus*, thereby reducing the risk of misidentification associated with phenotypic methods. Indeed, some *S. argenteus* isolates can only be distinguished from *S. aureus* through NRPS gene amplification. As such, these *S. argenteus* isolates could be misidentified as *S. aureus* if only the *nuc* gene is targeted ([13]).

A small proportion of isolates (3.03%) tested negative by NRPS-PCR. This slight discrepancy compared to *nuc*-based results may be due to intra-species variability. These cases could represent *S. aureus* genetic variants harboring mutations in the primer-binding regions, or possibly closely related species such as *S. argenteus*, which have already been identified in other studies ([20], [21], [22]).

Moreover, these NRPS-negative isolates highlight the need for more in-depth studies in Côte d’Ivoire, including sequencing-based approaches, to better understand the diversity of species within the *S. aureus* complex involved in human infections in the region.

No *S. schweitzeri* isolates (*nuc*-negative, NRPS size: 340 bp) were detected in this study. Therefore, no *S. schweitzeri* was misidentified as *S. aureus*. This result is consistent with that of Großmann et al. [8]. In their study, no *S. schweitzeri* was found in clinical isolates from Gabon either. Therefore, although this staphylococcal species occurs in African wildlife and exhibits many pathogenicity factors already known from *S. aureus*, it appears unable to cross the species barrier to humans ([17], [23]). One reason could be the absence of factors that enable *S. schweitzeri* to evade the human immune system ([14]). In addition, reported human cases to date have involved colonization in individuals likely to have had direct contact with wild animals, conditions that do not reflect the epidemiological profile of patients treated at the University Hospital of Angré.

Although no human infections have been observed so far, the potential for *S. schweitzeri* to overcome the host barrier through prophage acquisition warrants attention. In *S. aureus,* for example, immune evasion genes like *scn* and *sak* are acquired via the φSa3 prophage, which plays a crucial role in host adaptation and immune system evasion [24]. Given the close phylogenetic relationship between *S. schweitzeri* and *S. aureus*, it is surprising that φSa3 has not yet been identified in *S. schweitzeri* [8]. Continuous genomic surveillance and ongoing monitoring of patients are essential to determine whether *S. schweitzeri* will appear in clinical settings in the future – an indication that the species barrier may have been overcome. The One Health approach emphasizes the interconnectedness of human, animal, and environmental health, highlighting the need for a comprehensive strategy in monitoring zoonotic risks. As human encroachment into wildlife habitats increases, the likelihood of zoonotic spillover events and the emergence of novel infectious diseases grows, further underscoring the importance of continued vigilance.

### Evolution of the prevalence of MRSA in Côte d’Ivoire and comparison with previous data

The results of this study indicate a high prevalence of MRSA strains. The cefoxitin test identified 50.3% of isolates as resistant, and among them, over 80,12% carried the *mecA* gene. This rate is significantly higher than values reported in previous studies in Côte d’Ivoire: A study by Krizo et al. [25], conducted at the Pasteur Institute of Côte d’Ivoire, found a 41.9% MRSA rate among approximately thirty clinical isolates analyzed. Another study conducted between 1998 and 2001 in Abidjan reported a 25% MRSA rate among hospital isolates, with these strains primarily found in blood cultures and purulent secretions [26].

Compared to these earlier studies, our analysis reveals a significant increase in MRSA prevalence, with rates rising from 25% in the early 2000s to 41% in 2020 and 50% in 2025. This rapid increase indicates a continuous progression of MRSA infections and an ongoing rise in resistance to β-lactams in Côte d’Ivoire, representing a serious threat to the management of bacterial infections.

Globally, the WHO GLASS 2022 report indicates that the median MRSA rate in countries with effective surveillance systems is 6.8%, with a global average of 35% [27]. Our results show that Côte d’Ivoire far exceeds these reference values, highlighting a major antibiotic resistance issue in the country.

However, a limitation of our study is the lack of information on patients’ medical histories, such as recent antibiotic use and the duration of hospitalization. These factors are crucial for understanding the origin and high prevalence of the identified MRSA strains. Due to the lack of this data, it is possible that other factors, notably community transmission. Several factors such as unsupervised self-medication, residential promiscuity, and poor hygiene conditions, are associated with this community transmission of MRSA [28, 29]. Moreover, it is also important to note that the area where the University Hospital of Angre was built less than 10 years ago was a sparsely urbanized vegetation zone. Recent urbanization has led to a rapid increase in the population without necessarily having health infrastructure keep pace at the same rate. These conditions could play a role in the high prevalence of MRSA in this study. The analysis of the factors associated with resistance revealed a significant link with the hospital department from which the samples originated, particularly the surgical, adult medicine, and intensive care services. These services appear to play an important role in the spread of MRSA, likely due to more frequent antibiotic use or the increased fragility of the patients being treated there. A study conducted in Cameroon also showed that nasal colonization by MRSA significantly increased during hospitalization in surgery, rising from 13.5% at admission to 30.8% at discharge, which clearly illustrates the impact of the service on transmission [30]. In contrast, we did not observe any association with the type of sample. Among children, there is a higher proportion of methicillin-sensitive strains (MSSA), but they still remain the group with the largest number of MRSA strains in absolute value. This can be explained by their strong representation in the overall sample, but also by early exposure to antibiotics, whether in the community or hospital settings. Moreover, age seems to be an important factor: older subjects were more often carriers of MRSA, which has already been described in other studies [31, 32]. This distribution may be explained by the greater vulnerability of populations to infections and their more frequent exposure to healthcare settings, which are well-known factors for MRSA transmission [33].

Historically associated with healthcare-associated infections, MRSA is now increasingly found in the community. For instance, in Quebec, Canada, the proportion of community-acquired MRSA strains rose from 19% in 2016-2017 to 46.5% in 2019-2020, representing an increase of over 145% [34]. This evolution reflects the growing spread of these bacteria in the community

In this context, it is essential to strengthen the surveillance of these bacterial infections in Côte d’Ivoire, with a focus on vulnerable populations. Improved monitoring of clinical data would also help establish more precise links between infections and patients’ health status. Additionally, measures to promote hygiene and the rational and appropriate use of antibiotics are crucial to limit the spread of these resistant strains.

## Conclusion

This study shows a high prevalence of *S. aureus* in humans, confirmed in over 96% of cases through targeted genes, *nuc* and NRPS. No other species from the S. *aureus* complex were detected. More than half of the isolates exhibit methicillin resistance, with 50.3% of isolates being resistant, and an increase in MRSA prevalence with age. These data highlight the importance of antibiotic resistance surveillance and the contribution of molecular solutions for better identification of *S. aureus* and closely related species. Further studies, including diverse sampling that takes into account both clinical and carriage samples from different human and animal populations, along with sequencing analyses, will be necessary to better characterize the diversity and distribution of *S. aureus* complex species in Côte d’Ivoire.

## Data Availability

All relevant data are included within the manuscript. The full raw dataset, consisting of an Excel file with data from samples, is available from the corresponding author upon reasonable request.

## Acknowledgments

We would like to express our gratitude to everyone who contributed to the publication of this study.

We extend our sincere thanks to the Angré University Hospital, particularly the medical biology laboratory and the bacteriology unit, for their invaluable support, as well as for providing the biological materials and equipment necessary for this study.

Our appreciation also goes to the Pr. Koffi Stéphane, Pr. Gonedele Sery and Dr. Pakora from Treichville University Hospital and the UFR Biosciences, for their excellent field infrastructure and support in molecular biology work.

We also thank Institut für Medizinische Mikrobiologie, Universitätsklinikum Münster, Germany for providing the reference strain of *S. schweitzeri*.

## Contributor Role

**Conceptualization**: Kacou-Ndouba Adele, Djaman A Joseph, Niemann Silke

**Data Curation**: Obouayeba Djaman Nguessan Carole (ODNC)

**Formal Analysis**: Obouayeba Djaman Nguessan Carole, Kofi Kobina Amandze Adams

**Resources**: Coulibaly-Diallo G M, Bahan Armel-Joel, Kacou-Ndouba Adele, Niemann Silke

**Investigation**: Obouayeba Djaman Nguessan Carole, Bahan Joel Armel

**Methodology**: Obouayeba D N Carole, Niemann Silke, Coulibaly Edwige

**Supervision**: Djaman A Joseph, Kacou-Ndouba Adele

**Validation**: Obouayeba D N C, Niemann Silke, Kacou-Ndouba Adele, Djaman A Joseph

**Visualization**: Obouayeba D N Carole, Kofi Kobina Amandze Adams

**Writing – Original Draft Preparation**: Obouayeba Djaman Nguessan Carole

**Writing – Review & Editing**: Obouayeba D N Carole, Niemann Silke, Coulibaly Gniwele Edwige,Bahan Armel Joel, Djaman A Joseph, Kacou-Ndouba Adele

All authors read and approved the final manuscript.

